# Deep-learning Segmentation of Pediatric Brain Tumors using Ratio Maps of T1w/T2w MRI Signal Intensity

**DOI:** 10.1101/2025.04.09.25325530

**Authors:** Daniel Griffiths-King, Timothy Mulvany, Heather Rose, Jan Novak

**Affiliations:** Aston Institute of Health and Neurodevelopment, College of Health and Life Sciences, Aston University, Birmingham, UK; Medical Physics and Clinical Engineering, Nottingham University Hospitals NHS Trust, Nottingham, UK; Institute of Cancer and Genomic Sciences, University of Birmingham, Birmingham, United Kingdom

**Keywords:** Pediatric, Brain Tumor, Neuro-Oncology, Segmentation, BraTS Challenge

## Abstract

**Introduction:** T1w/T2w ratio mapping, combining voxel-wise signal intensities in T1-weighted (T1w) and T2-weighted (T2w) structural MRI, has been used to investigate cortical architecture in the brain, but has also shown promise in tissue discrimination, even in tumor tissue.

**Methods:** The current study aimed to investigate whether the inclusion of these established T1w/T2w ratio maps, or a similar T1w – T2w combined map, can improve performance on a novel task; automated segmentation of tumor tissue in pediatric brain tumor cases from the BraTS-PED 2024 dataset.

**Results:** We demonstrate that T1w/T2w ratio maps do not improve deep learning models for segmenting tumor subregions using nnU-Net.

**Conclusions:** Overall, our results suggest that including well-established combinations of T1 & T2w imaging modalities in addition to the already utilized mpMRI, as a potential method of input data augmentation, does not provide added value in the task of segmentation of pediatric brain tumors.

**Summary Statement:** The combination of T1w and T2w MRI images of the brain serves as a method to derive additional inputs for deep learning models. Their inclusion does not improve state-of-the art automated segmentation of pediatric brain tumors from standard MRI modalities.

**Key Results:** • Baseline segmentation differed for different tumor subregions. • Segmentation performance when T1w/T2w ratio maps were included, was similar to baseline for all tumor subregions. • Accuracy of enhancing tumor had greatest increase through inclusion of T1w/T2w Ratio Map but was not a statistically significant improvement.

## I. Introduction

MRI is essential for assessing pediatric brain tumors. Beyond traditional reporting, quantitative MRI analysis identifies in-vivo biomarkers to support diagnosis, predict histopathological status, assess treatment response, and predict prognosis [1]. Accurate tumor boundary delineation is required to quantify biomarkers. To avoid manual, labor-intensive methods, automated MRI segmentation via deep learning has gained significant traction.

Typically, automatic segmentation utilizes clinically acquired MRI including pre/post contrast T1-weighted (T1w & T1w-CE), T2-weighted (T2w), and T2 Fluid Attenuated Inversion Recovery (T2-FLAIR). This study introduces the T1w/T2w ratio map as a novel model input. Combining T1w and T2w images via a pixel-wise ratio, T1w/T2w mapping provides a high-resolution, non-invasive measure of cortical architecture, distinguishes cortical areas, and minimizes shared field inhomogeneities [2-4].

Historically, T1w/T2w mapping has been an in-vivo proxy for myelin content [2] and approximates T1w and T2w relaxometry (R1 & R2 rates) in clinical scenarios where quantitative MRI is impractical [5]. Prior research links T1w/T2w ratio with R1/R2 values in tumors [6, 7], and can identify non-enhancing regions in glioma [7] – an area where pediatric segmentation methods often struggle [8].

Utilizing T1w/T2w ratio maps for automated segmentation may improve tissue discriminability [9], as shown between healthy and glioma tissue for threshold-based segmentation [10]. Similarly, combined T1w-T2w maps, employing pixel-wise scaling rather than strict ratios, outperform individual T1w and T2w MRI in deep-learning segmentation of the claustrum [11]. A combination of existing inputs into deep-learning models, these ratio/combination maps represent a method for data extension to extract additional imaging features.

This exploratory study evaluates T1w/T2w ratio and combined maps as a novel input to nnU-Net, a leading deep-learning segmentation framework with proven efficacy in brain tumor segmentation [12, 13]. nnU-Net automatically adapts preprocessing, network architecture, training, and post-processing, in response to the training data [12]. It is hypothesized that incorporating T1w/T2w maps will improve accuracy of automatic segmentation of pediatric brain tumors using the Brain Tumor Segmentation Pediatrics Challenge (BraTS-PED) 2024 dataset [8].

## 2. Materials and Methods

### 2.1 Data

#### Participants

The CBTN-CONNECT-DIPGR-ASNR-MICCAI BraTS-PEDs 2024 Challenge dataset is a retrospective cohort consisting of data from n=464 pediatric patients with high-grade glioma (e.g. high-grade astrocytoma, DMG and DIPG). Only data from the training (n=261) cohort, where both the MRI and training labels are available, is used in the current study, due to access restrictions on validation/testing cohorts. Further details are published elsewhere [8]. Data were obtained through Synapse and MedPerf systems [14] (ID syn511569100029) and Aston University College of Health and Life Sciences Research Ethics Committee (#HLS21041) granted ethical approval for secondary analysis. Data was accessed in May 2024, and authors had no access to identifying data.

#### MRI

The BraTS-PEDs dataset contains multiparametric MRI (mpMRI) sequences; T1w, T1w-CE, T2w, and T2-FLAIR. Data was already pre-processed, using the “BraTS Pipeline” (through the Cancer Imaging Phenomics Toolkit (CaPTk) and Federated Tumor Segmentation (FeTS) tool), and anonymized - removing protected DICOM headers and MRI defacing [8].

#### Data Annotations – Tumor Sub-regions

Reference annotations of four tumor subregions are provided for the training cohort: “enhancing tumor” (ET), “non-enhancing tumor” (NET), “cystic component” (CC) and “edema” (ED). Two additional labels are generated through combinations of subregions, “tumor core” (TC) combining ET, NET, and CC, and “whole tumor” (WT) - the entire tumorous region combining ET, NET, CC and ED. Generating reference annotations involved semi-automated segmentation, iterative refinement/editing of labels, and final review by neuroradiologists [8]) .

#### Generating T1w/T2w Maps

##### T1w/T2w Ratio Map

Ratio maps were calculated through normalization/standardization and straightforward division of T1w by T2w images. These were calculated as:

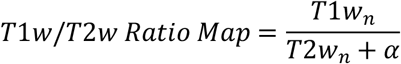

where *T*1*w*_*n*_ & *T*2*w*_*n*_ are normalized T1w & T2w images (normalization described in supplementary materials) and *α*=0.001 (to prevent null division). This follows guidance in [15].

##### Combined T1w-T2w Map

The T1w-T2w combined map was calculated (voxel-wise) as:

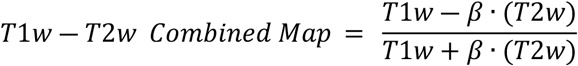

where *β* is a scaling factor for the purposes of normalization (described in supplementary materials). This echoes the approach in [11]. These were termed a ‘combined’ map as they are not strictly a ratio image.

### 2.2 Model Architectures

The nnU-Net residual encoder variant was used as the benchmarking model^1^ [16]. This uses residual blocks in the encoder, which show benefit in brain tumor segmentation tasks [16], where the convolution block’s input is added to the output, preserving information from previous layers.

This study tests three configurations of model inputs; a) a baseline model using the four original mpMRI scans (T1w, T1w-CE, T2-FLAIR and T2w), b) where the T1w/T2w ratio or c) T1w-T2w combined map is included as an additional 5^th^ input channel (see Figure 1).

**Figure 1.**
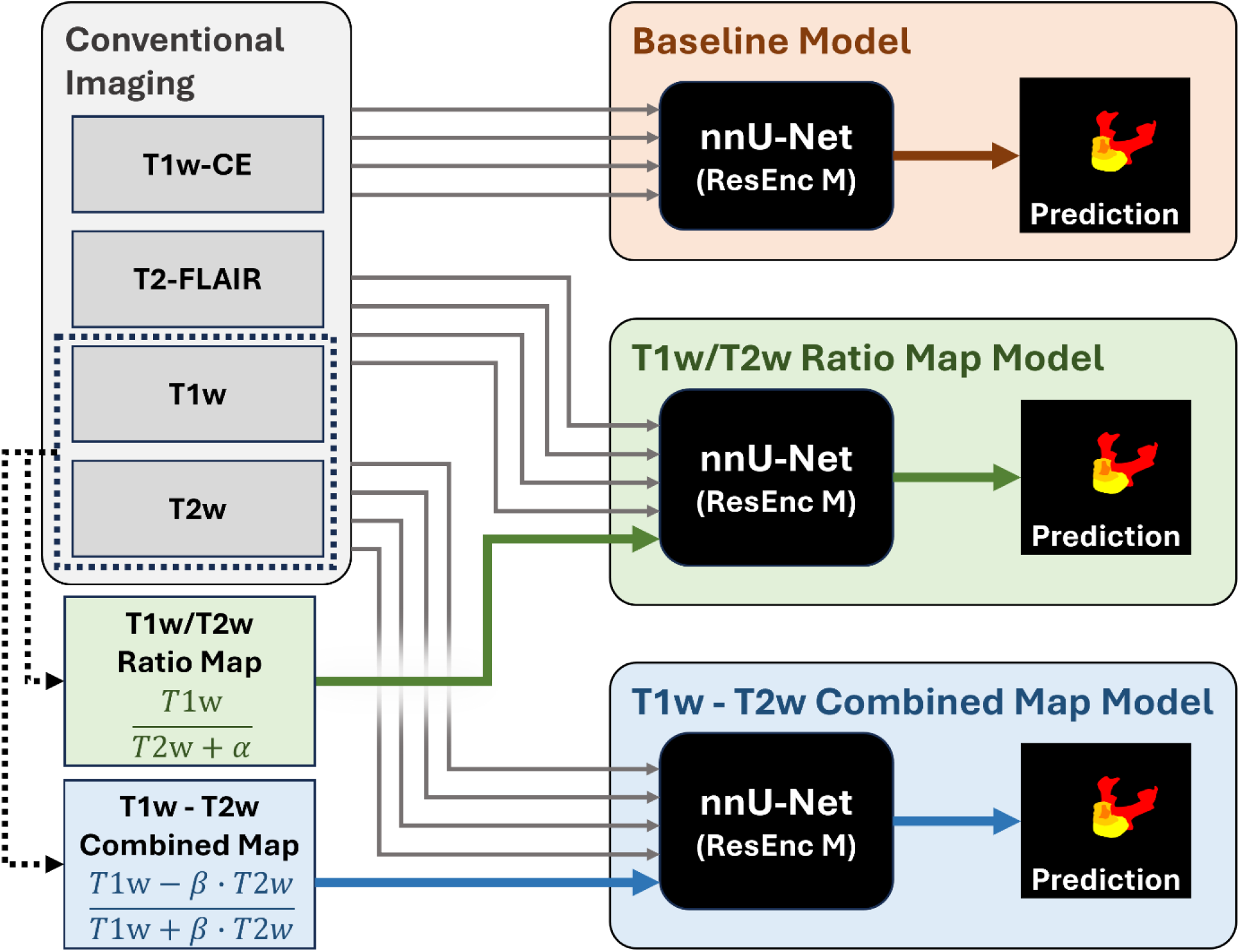
Workflow of the models tested in the current study, including generation of Ratio and Combined maps.

### 2.3 Training and Evaluation

Training followed nnU-Net’s default methodology, including on-the-fly data augmentation [12], and 5-fold internal-validation with subject-level splits. Each run lasted 100 epochs with a batch size of 2. Networks were optimized using stochastic gradient descent with Nesterov momentum=0.99, initial learning rate=0.01 and polynomial decay schedule. Models from each fold were ensembled for evaluation per nnU-Net’s default behavior. Reported results reflect performance on the held-out fold during validation.

Performance was evaluated using Dice score (DSC) [8], measuring overlap between automated segmentation and reference labels across the four tumor subregions. To assess segmentation improvement over the baseline model (standard clinical imaging modalities), we compared models additionally incorporating the combined or ratio maps, using a repeated-measures, one-tailed t-test. A Bonferroni corrected α_crit_=0.0125 addressed multiple comparisons over the 4 tumor subregions. For non-normally distributed Dice scores, Wilcoxon signed rank tests were adopted.

#### Computational Resources

Experiments were conducted with PyTorch (v2.3.0 + CU12.1) on two NVIDIA Quadro RTX6000 GPUs with 24GB VRAM. Ensembling of predictions from differing folds was performed using Python 3.9.19 & NiBabel 5.2.1.

Additional analyzes are reported in supplementary materials.

## 3. Results

Table 1 presents the segmentation performance accuracy across models and tumor subregions.

**Table 1.** Results of segmentation performance for each model during internal validation, across tumor subregion labels.

| Model | Dice Score |  |  |  |  |  |  |  |  |  |  |  |
| --- | --- | --- | --- | --- | --- | --- | --- | --- | --- | --- | --- | --- |
|  | ET |  |  | NET |  |  | CC |  |  | ED |  |  |
|  | Mean | SD | Med. | Mean | SD | Med. | Mean | SD | Med. | Mean | SD | Med. |
| Baseline | 0.550 | 0.350 | 0.697 | 0.783 | 0.233 | 0.871 | 0.271 | 0.334 | 0.020 | 0.174 | 0.267 | 0.000 |
| T1w/T2w Ratio Map | 0.569 | 0.344 | 0.703 | 0.773 | 0.250 | 0.880 | 0.272 | 0.325 | 0.077 | 0.160 | 0.254 | 0.000 |
| Combined T1w-T2w Map | 0.533 | 0.346 | 0.689 | 0.785 | 0.227 | 0.873 | 0.266 | 0.332 | 0.005 | 0.158 | 0.256 | 0.000 |

Note. T1w = T1-weighted MRI, T2w = T2-weighted MRI, ET = Enhancing Tumor, NET = Non-enhancing Tumor, CC = Cystic Component, ED = Edema, Med. = Median

### Baseline Model

Performance of the baseline model during internal validation (cross-fold) was highest for the NET label 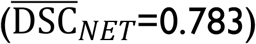, compared to ET 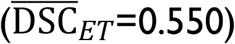. Performance was lowest for the CC and ED subregions 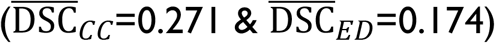.

### T1w/T2w Ratio Map Model

Performance of the model when T1w/T2w ratio maps were included, was similar to baseline for all tumor subregions during internal validation (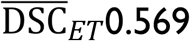 vs. 0.550, 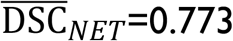 vs. 0.783, 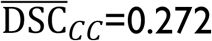 vs. 0.271, 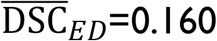 vs. 0.174, for ratio map versus baseline models respectively). Accuracy of ET segmentation had the largest increase (+ 0.019) across all tumor subregions and all included models when tested against the baseline model but still did not reach statistical significance. No change in Dice between baseline and the ratio map models was observed to be statistically significant across the tumor subregions (all p > 0.0125).

### Combined T1w-T2w Map

When the T1w-T2w combined map was included performance was similar to the baseline model for all tumor subregions during internal validation (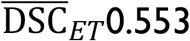 vs. 0.550, 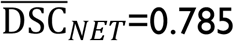 vs. 0.783, 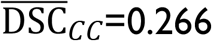 vs. 0.271, 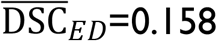 vs. 0.174, for ratio map versus baseline models respectively). Whilst ET & NET delineation was marginally improved, larger drops in CC and ED segmentations were observed. No change in Dice between baseline and combine map models was statistically significant (all p > 0.0125).

## 4. Discussion

This study tested whether adding T1/T2 ratio maps improved tumor segmentation in a large, well annotated, pediatric brain tumor MRI dataset. Despite prior literature suggesting potential added benefit, the results showed no significant increase in segmentation performance for ratio or combined approaches.

Previous studies have identified group-level differences in mean T1w/T2w ratio across various pathologies, including adenohypophyseal tumors [17], lung cancer [18] and cerebellar-subtype multiple systems atrophy [19]. Moreover, T1w/T2w ratio as a surrogate for MR relaxometry, had reported benefits in discriminating healthy and tumorous brain tissue [7, 9, 10]. This made it a good target for inclusion as an additional input modality for automated tumor segmentation. In this study, inclusion of T1w/T2w ratio maps, alongside standard mpMRI modalities, did not increase segmentation accuracy. Differences in Dice for specific labels were not statistically significant, and overall, performance on the dataset was low compared to published benchmarks (e.g. BraTS-PEDs 2023 [20]). The interpretation of these findings is that ratio maps do not appear to add any significant additional information, not ascertained through simple additive convolutional differences and the additional computational burden required to generate the maps is not justified.

Previous work which highlighted the benefit of T1w/T2w ratio maps in improved segmentation did not assess the segmentation improvement of including the T1w/T2w ratio maps on top of the existing mpMRI approaches, instead directly comparing to either a T1w only, or T2w only model [11]. Using a single modality baseline models does not allow us to assess whether segmentation improvement is solely due to the inclusion of ratio map inclusion or additional information from the second, T1w or T2w, modality [21].

This approach was explored during the development phase of the BraTS-PED 2024 challenge however, it was ultimately abandoned for more effective alternatives. In reporting these null findings, we hope to reduce potential duplication of effort in future challenge contexts and future research efforts by reducing the file-drawer-problem in the medical-imaging field [21].

Overall, combining T1w/T2w imaging modalities does not appear to add value for pediatric brain tumor segmentation when integrated with mpMRI inputs.

## Data Availability

This analysis is a secondary analysis of publically available data. The data originated from the BraTS-PED 2024 challenge, controlled, and accessed via through Synapse project (ID syn51156910) as stated and cited in the manuscript.

https://www.synapse.org/Synapse:syn53708249/wiki/626323

## Funding Acknowledgement

Thanks to Help Harry Help Others for funding TM via a PhD Studentship. DGK was funded by Aston University College of Health and Life Sciences via a post-doctoral award to DGK and JN.

## Footnotes

1 As per https://github.com/MIC-DKFZ/nnUNet/blob/96253e9dae2e7879520ab5dfc1c84aefe0f818e7/documentation/resenc_presets.md

